# “The further along I get the more I feel like growth is possible”: lived-experience narratives of the shape of recovery in borderline personality disorder

**DOI:** 10.64898/2026.10.08.26365022

**Authors:** Verenice Torres, Thea M. McAfee, Paulina Tein, Alexandra A. Alario, Wenjing Luo, Kaylin Foss, Sarah K. Fineberg, Sharon M. Nelson

**Author notes:** Corresponding author, (VT). These authors contributed equally to this work (Verenice Torres, Thea M. McAfee).

## Abstract

Borderline personality disorder (BPD) has traditionally been viewed as one of the most chronic psychiatric disorders. Although longitudinal studies have demonstrated promising rates of symptomatic remission, recovery and other clinical outcome measures often remain comparatively low. Qualitative research offers an important perspective on how individuals with BPD may extend a more representative view of improvement and change. We collected narratives from 41 participants with a self-reported diagnosis of BPD and analyzed them using thematic analysis. Its national, virtual recruitment approach enabled a relatively large qualitative sample diverse in gender identity, age, U.S. geographic region, treatment history, and current clinical severity, extending the range of perspectives represented in prior work. Three central themes were identified. The first, *Differing attitudes toward language*, reflected participants’ experiences of recovery language as paradoxically both validating and stigmatizing. While individual language preferences varied, participants generally converged on a shared understanding of what recovery meant to them. The second, *Recovery as a dynamic and relational process*, described recovery as nonlinear, effortful, and closely tied to interpersonal experiences. The third, *Crossroads and uneven change over time*, highlighted the role of personal agency, critical decision points in seeking treatment, and the persistence of internal distress even as behavioral symptoms improve. Findings from this study both replicate and extend existing qualitative research by discussing how individuals with BPD understand recovery in ways that may not be captured by traditional clinical outcome measures. These results emphasize the importance of incorporating lived-experience perspectives when defining recovery and setting treatment goals.

## Introduction

Borderline personality disorder (BPD) is a serious psychiatric condition that has profound impacts on quality of life, productivity, and lifespan. It is common (1-5.9% prevalence [1]), accounts for up to 25% of required inpatient psychiatric care in the United States [2, 3], and is associated with markedly elevated rates of medical disability and mortality [4, 5].

BPD is characterized by distressing experiences of affective lability, intense emotional reactions, and unstable relationships [6]. Access to high-quality care is limited by small numbers of trained providers and widespread stigma in the healthcare system, including beliefs that remission is not possible for individuals with BPD [7]. BPD has traditionally been viewed as chronic and intractable [1]. However, over the past 25 years, a growing literature has challenged this prognostic pessimism by demonstrating reduction in number and intensity of BPD symptoms over time [1, 8, 9].

However, methods focused on counting symptoms to quantify change may not fully capture all dimensions of an individual’s experience of changes in BPD severity over time. Limitations in the clinical conceptualization of recovery for people with mental health problems have led to articulation of subtypes of recovery: clinical (e.g., improvement in symptom severity), functional (e.g., work, school, relational roles) and personal (e.g., progress toward one’s own hopes and goals) [10, 11]. The narrow conceptualization of clinical outcome in the existing literature, challenged by evidence suggesting that remission is an insufficient indicator of recovery, complicate efforts in meaningfully characterizing the course of BPD in a way that accurately reflects lived experiences. A fuller picture of the functional outcome of BPD may be achieved by recognizing the limitations of standardized quantitative approaches to measuring the full breadth of recovery and emphasizing more qualitative ones.

Over the past fifteen years, several qualitative studies describing the experiences of recovery in BPD have been published [12–23]. Past work has utilized a variety of qualitative methods to explore potential differences between clinical perspectives and lived experience [24, 25], including thematic analyses of interviews [22, 26, 27] and creative methods like analysis of participant-created visual works [14, 28, 29], and Instagram posts [18].Most of the work focused on patients currently enrolled in specialty personality disorder services, though Ng et al. (2019) interviewed a community sample of 14 people (seven in recovery) about recovery in BPD, Esposito et al. (2021) reviewed Instagram posts on 14 accounts where the posts identified the writer as having BPD, and Liljedahl et al. (2023) interviewed nine women with who identified as in recovery for at least two years from BPD and severe self-injury about their views on recovery and life worth living. Samples were predominantly very small, though six had 10-12 participants, and two included 48 individuals. Samples were primarily (>80%) or entirely women and most were drawn from single treatment sites or geographic regions.

This previous work emphasized problems with the term recovery [13–15, 22], the dynamic nature of the recovery process [12–15, 19], and the role of social relationships [12–18, 20]. Nearly all the papers included themes about views of self and identity, and some discussed symptoms (emotion dysregulation, suicidality, non-suicidal self-injury). Several papers discussed the importance of aspects of functional recovery [13, 14, 16, 20]. Less common themes included direct discussion about clashes between clinical and personal recovery goals, challenges in treatment, belief systems (meaning, gratitude, faith), and maintaining self-care. Perhaps surprisingly, direct focus on stigma was also rare.

In this manuscript, we report on a qualitative analysis of 41 interviews about the nature of recovery with people with lived experience of BPD. We sought to replicate and extend prior work with a large cohort including adults with carefully characterized current and past BPD, diverse histories of treatment, age, gender identity, and geography. We leveraged this relatively large qualitative sample to identify three overarching themes and describe several important areas of tension in the lived experience of change over time in BPD. Given the rapidly changing landscape in which BPD-related information, diagnostic disclosure, and treatment are ever more readily accessible, we were particularly interested in how people who have been affected by BPD think about the term recovery, about factors that have helped and hindered their recovery, and how they thought about themselves and their relationships.

Existing literature provides a valuable holistic overview of lived-experience perspectives of recovery, an overview which the present study aims to expand upon, drawing upon our large, heterogenous sample to further investigate topics like components of effective/ineffective treatments [24, 27, 30, 31], the central role of interpersonal relationships in the recovery journey [14, 16, 26, 32], and the nonlinear nature of recovery [32, 33] with greater specificity. The present study seeks to delve deeper into these central topics of language and meaning-making, examining how lived-experience perspectives on recovery and BPD have (or have not) shifted since the prior reports. In addition, this study aims to address gaps through recruiting a large sample diverse in gender identity, age, geographic region, and current clinical severity than many of the existing studies.

## Method

The study reporting is informed by the Standards for Reporting Qualitative Research.

### Participants, Sampling, and Recruitment

#### Ethics

The procedures for this study were reviewed and approved by the Yale University Institutional Review Board (HIC# 2000026215). Study procedures were conducted in accordance with the Declaration of Helsinki. This study was not pre-registered.

#### Recruitment

Individuals were recruited via social media (Instagram, Facebook) throughout the United States as part of a large virtual study. Potential participants were screened through a brief phone call then a Zoom visit for study eligibility. Participants who self-reported having received a BPD diagnosis from a mental health professional, were fluent in English, and were over 18 years old were included in the present analysis. Participants were excluded if there was indication of any neurodevelopmental or primary psychotic disorder, severe substance abuse, or a history of neurological or learning problems.

#### Participant characteristics

The sample size (*n* = 41) and breadth of recruitment allowed us to capture a diversity of demographic representation and experiences with regards to BPD treatment and lived experience (see Tables 1-3). Our sample included a broad representation of gender identities (73% women, 10% men, 17% nonbinary or genderqueer), expanding on many qualitative studies on BPD which focus solely on women’s experiences [14, 22, 26, 34].

**Table 1.** Participant Demographics.

|  | **Total (N=41)** | **Current BPD (N=29)** | **Past BPD (N=12)** |
| --- | --- | --- | --- |
| **Gender** |  |  |  |
| Female | 30 (73.2%) | 21 (72.4%) | 9 (75.0%) |
| Male | 4 (9.8%) | 2 (6.9%) | 2 (16.7%) |
| Nonbinary/genderqueer | 7 (17.1%) | 6 (20.7%) | 1 (8.3%) |
| **Age** |  |  |  |
| Mean (SD) [Min, Max] | 27.5 (7.58) [18.0, 48.0] | 26.1 (7.26) [1.0, 48.0] | 30.8 (7.63) [20.0, 43.0] |
| **Race** |  |  |  |
| Native American | 0 (0%) | 0 (0%) | 0 (0%) |
| Asian | 6 (14.6%) | 4 (13.8%) | 2 (16.7%) |
| Black | 1 (2.4%) | 1 (3.4%) | 0 (0%) |
| MENAA | 1 (2.4%) | 1 (3.4%) | 0 (0%) |
| Pacific Islander | 3 (7.3%) | 1 (3.4%) | 2 (16.7%) |
| White | 28 (68.3%) | 20 (69.0%) | 8 (66.7%) |
| Multiracial | 2 (4.9%) | 2 (6.9%) | 0 (0%) |
| **Ethnicity** |  |  |  |
| Hispanic | 6 (14.6%) | 5 (17.2%) | 1 (8.3%) |
| non-Hispanic | 35 (85.4%) | 24 (82.8%) | 11 (91.7%) |

**Notes:** MENAA: Middle Eastern or North African

**Table 2.**
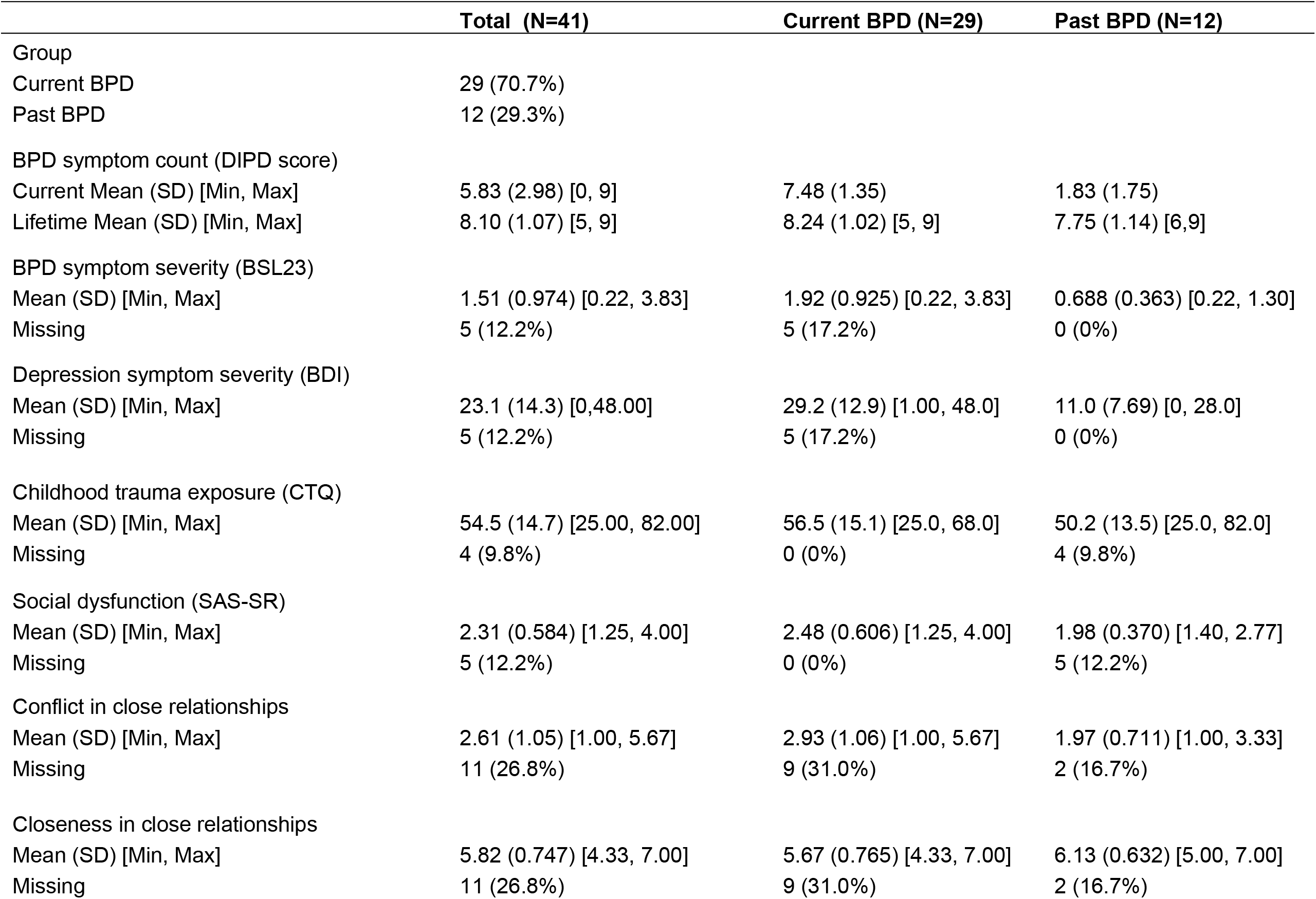
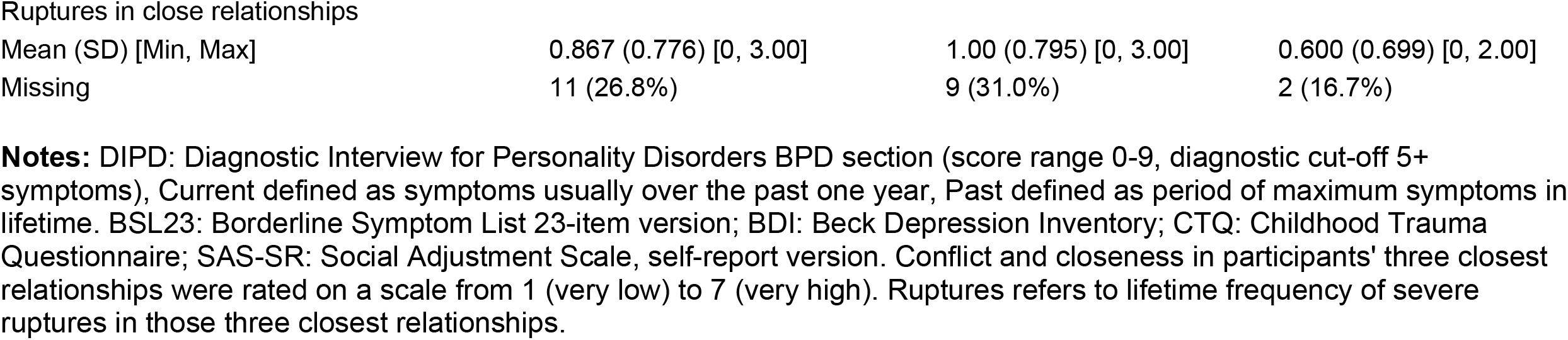
Participant Clinical and Social Characteristics.

|  | **Total (N=41)** | **Current BPD (N=29)** | **Past BPD (N=12)** |
| --- | --- | --- | --- |
| Group |  |  |  |
| Current BPD | 29 (70.7%) |  |  |
| Past BPD | 12 (29.3%) |  |  |
| BPD symptom count (DIPD score) |  |  |  |
| Current Mean (SD) [Min, Max] | 5.83 (2.98) [0, 9] | 7.48 (1.35) | 1.83 (1.75) |
| Lifetime Mean (SD) [Min, Max] | 8.10 (1.07) [5, 9] | 8.24 (1.02) [5, 9] | 7.75 (1.14) [6, 9] |
| BPD symptom severity (BSL23) |  |  |  |
| Mean (SD) [Min, Max] | 1.51 (0.974) [0.22, 3.83] | 1.92 (0.925) [0.22, 3.83] | 0.688 (0.363) [0.22, 1.30] |
| Missing | 5 (12.2%) | 5 (17.2%) | 0 (0%) |
| Depression symptom severity (BDI) |  |  |  |
| Mean (SD) [Min, Max] | 23.1 (14.3) [0, 48.00] | 29.2 (12.9) [1.00, 48.0] | 11.0 (7.69) [0, 28.0] |
| Missing | 5 (12.2%) | 5 (17.2%) | 0 (0%) |
| Childhood trauma exposure (CTQ) |  |  |  |
| Mean (SD) [Min, Max] | 54.5 (14.7) [25.00, 82.00] | 56.5 (15.1) [25.0, 68.0] | 50.2 (13.5) [25.0, 82.0] |
| Missing | 4 (9.8%) | 0 (0%) | 4 (9.8%) |
| Social dysfunction (SAS-SR) |  |  |  |
| Mean (SD) [Min, Max] | 2.31 (0.584) [1.25, 4.00] | 2.48 (0.606) [1.25, 4.00] | 1.98 (0.370) [1.40, 2.77] |
| Missing | 5 (12.2%) | 0 (0%) | 5 (12.2%) |
| Conflict in close relationships |  |  |  |
| Mean (SD) [Min, Max] | 2.61 (1.05) [1.00, 5.67] | 2.93 (1.06) [1.00, 5.67] | 1.97 (0.711) [1.00, 3.33] |
| Missing | 11 (26.8%) | 9 (31.0%) | 2 (16.7%) |
| Closeness in close relationships |  |  |  |
| Mean (SD) [Min, Max] | 5.82 (0.747) [4.33, 7.00] | 5.67 (0.765) [4.33, 7.00] | 6.13 (0.632) [5.00, 7.00] |
| Missing | 11 (26.8%) | 9 (31.0%) | 2 (16.7%) |
| Ruptures in close relationships |  |  |  |
| Mean (SD) [Min, Max] | 0.867 (0.776) [0, 3.00] | 1.00 (0.795) [0, 3.00] | 0.600 (0.699) [0, 2.00] |
| Missing | 11 (26.8%) | 9 (31.0%) | 2 (16.7%) |

**Notes:** DIPD: Diagnostic Interview for Personality Disorders BPD section (score range 0-9, diagnostic cut-off 5+ symptoms), Current defined as symptoms usually over the past one year, Past defined as period of maximum symptoms in lifetime. BSL23: Borderline Symptom List 23-item version; BDI: Beck Depression Inventory; CTQ: Childhood Trauma Questionnaire; SAS-SR: Social Adjustment Scale, self-report version. Conflict and closeness in participants' three closest relationships were rated on a scale from 1 (very low) to 7 (very high). Ruptures refers to lifetime frequency of severe ruptures in those three closest relationships.

**Table 3.**
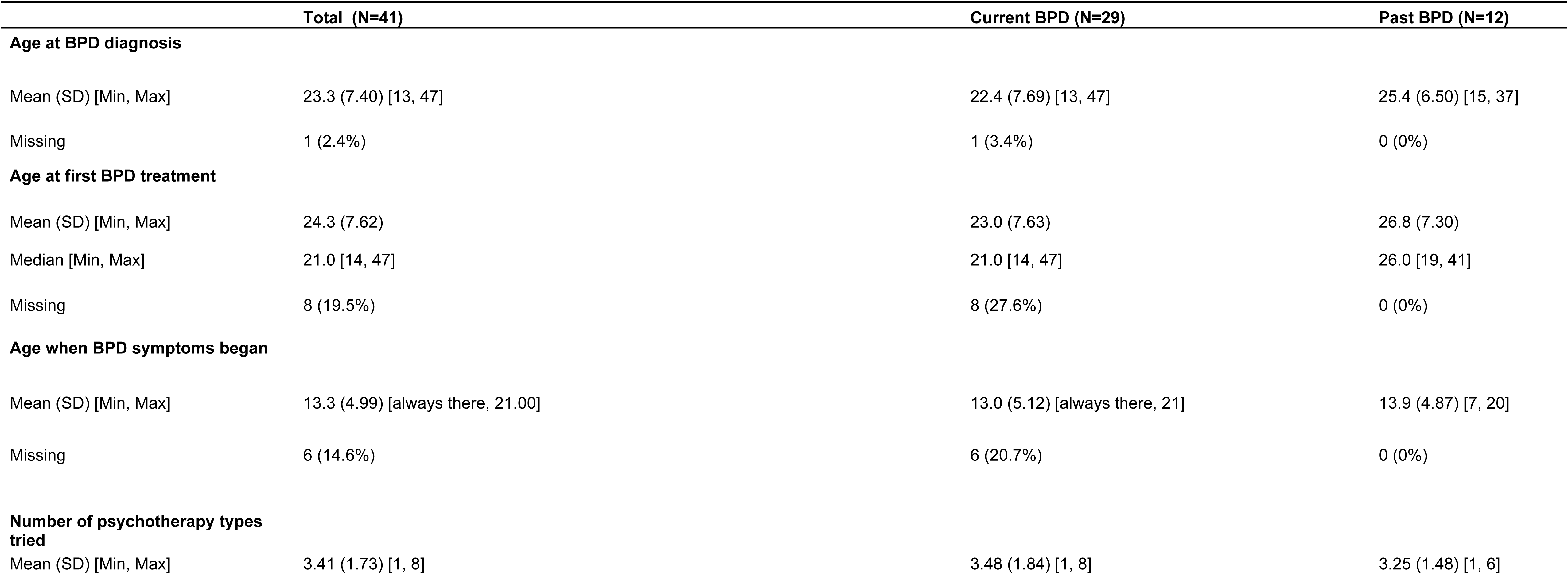
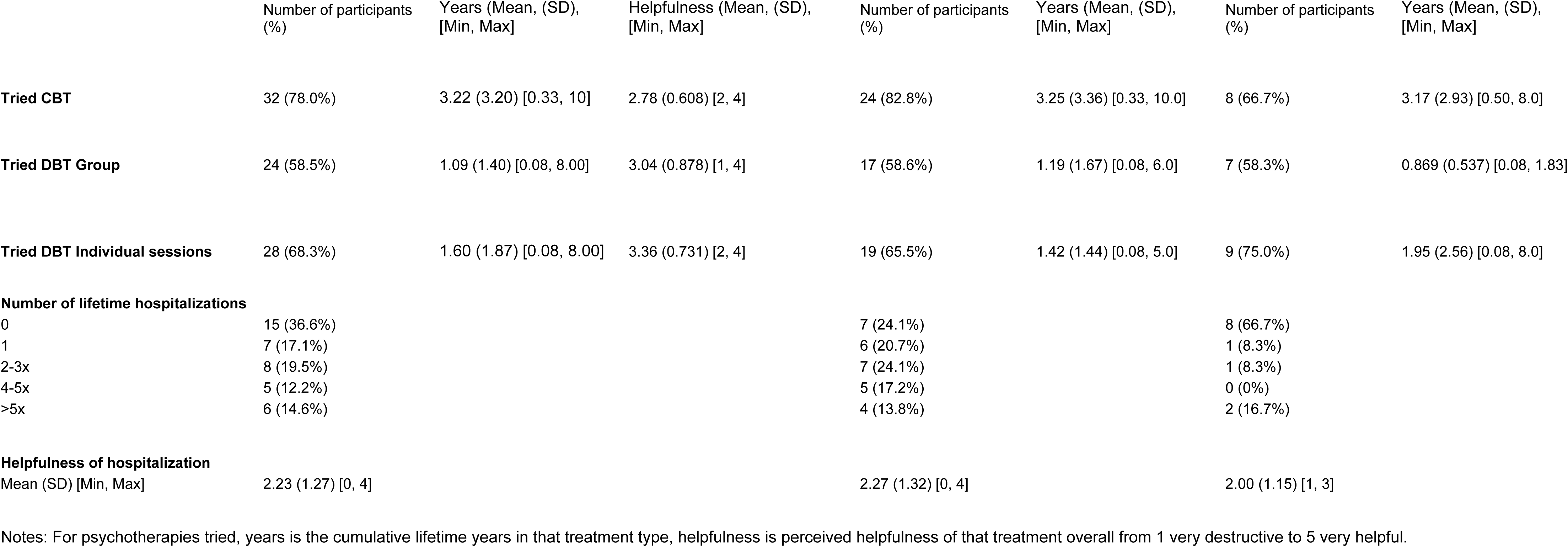
Participant Treatment History.

All participants completed an intake protocol consisting of a series of self-report measures and clinician administered interviews, including the Diagnostic Interview for DSM-IV Personality Disorders–Borderline Scale (DIPD-BPD). The DIPD-BPD consists of nine questions corresponding to the nine DSM-IV criteria for BPD, and each criterion is scored as present or not present [35]. Of the 41 participants included, 29 currently met diagnostic criteria (Mean DIPD-BPD = 7.48, SD = 1.35), while 12 last met criteria more than a year prior to the study (Mean DIPD-BPD = 1.83, SD = 1.75). Scores on the Borderline Symptom List – Short Form (BSL-23; [36, 37]), a measurement of current symptom severity, ranged from 0.217 (none or low) to 3.83 (extremely high, *M* = 1.51 (moderate), *SD* = 0.974). See Table 2.

Participants reported having tried multiple mental health treatments (*M* # of treatments tried = 3.41, [1–8], 78% tried cognitive behavioral therapy (CBT), 80.5% tried dialectical behavioral therapy (DBT)), with varying degrees of helpfulness (see **Table 3**).

### Study Design and Methodological Approach

#### Qualitative Interviews

Semi-structured interviews were conducted using an interview guide developed by VT based on a review of recovery literature and questions that arose in its reflective analysis (e.g., [13, 16, 38]). A phenomenological framework guided the study, with an emphasis on understanding recovery from the perspective of individuals with lived experience of BPD. Interview questions explored experiences of change, personal definitions of recovery, and factors that support or hinder recovery (see S1 Appendix). Open-ended questions were used to encourage detailed reflection and discussion. The guide was designed with the assumption that recovery is individualized and nonlinear [22, 33], with language selected to promote psychological safety and sensitivity to participants’ experiences. The interview guide was reviewed by study team members (SF and TM) and piloted with a colleague with lived experience of BPD, who provided feedback before interviews began.

Data were collected as individual semi-structured interviews in remote visits conducted by trained lab personnel. Zoom-generated interview transcripts were de-identified and proof-read by the coding team for accuracy, which began the data immersion process for coders not present for the interviews [39]. NVivo (Lumiverio, Version 13) was used for coding and analysis.

#### Visual representations of recovery

During interviews, participants were invited to draw visual representations of recovery using a collaborative digital canvas (e.g., Zoom Whiteboard, Google Jamboard). The unstructured format of this exercise was designed to facilitate creative expression and allow for additional approaches to expressing understandings of recovery.

### Thematic Analysis

Thematic analysis was selected due to its utility and previous use in exploring recovery in BPD [18, 22, 26, 27]. Coding followed a recursive process of inductive thematic analysis following the six phases recommended by Braun & Clark [39]: familiarization with the data, generating initial codes, searching for themes, reviewing themes, defining themes, and producing the report. [18, 22, 26, 27]

At each stage transcripts were coded by two separate coders, and discrepancies in rater codes were resolved through a team consensus process, following the framework outlined by Cofie et al. [40] to ensure high reliability while acknowledging the interpretivist epistemological paradigm of qualitative research. The coding team included two coders who were removed from data collection to address bias (AA and PT) and two coders with experience coding qualitative data (TM and VT). Another researcher with expertise in qualitative methods (SN) was consulted throughout the coding process.

## Results

This paper identified three central themes: 1) the role of language in recovery, 2) the nature of recovery, and 3) how the course of recovery is shaped. Some codes from the codebook were ultimately not included here, as they were outside the scope of this work. Themes and subthemes are illustrated with representative quotes in Table 4. Visual data from the drawing exercise were used to illustrate themes identified in the interviews, as shown in Fig 1.

**Table 4.** Themes and representative quotes.

|  |  |
| --- | --- |
| Theme 1: Differing attitudes toward language. |  |
| Subtheme 1a: Shared vocabulary: labels offer a framework for self and other. | Y2088: I kind of like that word [recovery] because it makes it easier to understand your own state. And I don't know, when I say that I'm in recovery from my mental health issues, it's just, I feel like I'm acknowledging the issues I had in the past and I'm also stating that I'm actively working on it. |
|  | Y1832: I think for me the word [recovery] has a deeper personal attachment to the word, like where you once lived in turmoil and now you no longer do so, you're in recovery, you know? I don't know, but I definitely relate to the word, and at least for me, I think it's a great way to describe where I'm at, because I'm not saying I'm recovered. That's the other thing. I'm not saying it's gone, like I'm healed, like I'm recovered. I'm feeling better and manageable. So I'm in this sort of recovery state. |
|  | Y2350: I feel like recovery is the [word] that makes the most sense logistically, because people understand what recovery is from substance use. And so, if we would use "decline in symptoms" or something, it's just not gonna take as much of a hold as recovery, you know, acknowledging that it's still there. |
| Subtheme 1b: "I'm not broken": stigma and black and white thinking. | Y1330: We don't really talk about people who have ADHD if they are medicated, and whatever, doing well, and don't have symptoms of ADHD. I don't think people call that recovery, or remission, because I feel like recovery needs to have the stigma of "broken" in order for it to make sense. And like, I don't know. I have friends with ADHD who would never say, "Oh, I've recovered from ADHD," because they have tools and medication that reduce their symptoms to the point where it doesn't affect them as much. I think maybe part of the reason why I hate the word recovery is because we don't use it for so many things that we could be using it for. And it's like, why is that? Why don't we say people recovered from ADHD? It's because we didn't think they were broken in the first place. |
|  | Y1612: I don't know that I would personally describe myself as in recovery for BPD. I would probably use the term "in treatment for". Because I guess recovery kind of gives the implication that, yeah, there's something wrong with you, or you're trying to fix something, whereas I don't think there's anything wrong with me. |
|  | Y1930: So definitely the way [recovery] was defined a lot of the time was no longer meeting criteria. So maybe you only have 3 instead of 5 symptoms. But for me, I feel like that's just super inaccurate and super dehumanizing because the human experience is so much more variable than that. |
| Subtheme 1c: Beyond labels: negotiating meaning and language of recovery | Y1930: I don't know that the word [recovery] itself is the problem. I think the conceptualization and definition for me is more of the issue. So the term in and of itself, I don't personally find issue with. It's really how medical professionals kind of conceptualize it and use it almost in place of remission, which I think is a little bit less colloquial, and a little bit less friendly to patients. |
|  | Y2045: Whatever's helpful to talk about the thing, and whatever other people use, I'll use it. |
|  | Y2575: I feel like this is a six of one, half dozen of the other kind of a situation, like, I don't have an issue with saying I'm "in remission" versus I'm "in recovery". But I can understand why people prefer to use the term "active recovery" versus "remission" ... I don't know that they're interchangeable, but I don't have an issue using them interchangeably... I think it's all about what a person is more comfortable using. |
| Theme 2: Recovery as a dynamic and relational process. |  |
| Subtheme 2a: "A lifelong journey": recovery as a process, not an endpoint. | Y1832: I think my personal belief is that once you have BPD, you can get into a state of recovery, but you'll always have some like – even if it's little triggers – you'll have stuff that you're kind of dealing with forever. You just get better at dealing with it. |
|  | Y2634: I feel like recovery is more of a term for physical illness, where you have a fever and you recover from a fever. You don't have that fever, you don't have the symptoms of the fever, whereas like a mental health thing, you could still have those symptoms, but you're better equipped to be able to go through life despite having the symptoms. |
|  | Y2692: I think obviously you can learn to avoid some of these harmful behaviors, and you can heal your relationships with people that have been affected by BPD. You could heal from some of the trauma that you know contributes to BPD happening... I certainly haven't been able to overcome all of the feelings that occur, the thoughts I have, the urges to do certain behaviors. But also, many aspects of my life have |
|  | improved, and I have learned to deal with those behaviors... but I wouldn't call that recovery, because recovery implies that everything is fixed... you just do the best you can. |
| Subtheme 2b: My recovery, my responsibility: embracing agency. | Y2649: I've done a lot of work to learn how to manage it but I would say, it's definitely still there. It's still like, I don't want to say an everyday battle, but you know, some days are worse than others. And so, I still have to actively put a lot of work and a lot of energy into not reacting like the way I would have before treatment. |
|  | Y2359: It's [recovery] more like there's not a full treatment that's just going to fix everything. It's like they have to take active steps in their life to be as okay as they can be. |
| Subtheme 2c: Healing and harm: connection as both struggle and goal. | Y2292: [BPD] is not super super impairing in my life by any means, and I think that it really only shows up in very certain areas of my life, like specifically with my partner. That's where everything comes out, and it's kind of a disaster. |
|  | Y1930: My relationships were always out of whack... I had a habit of choosing partners who were not excellent... The relationships were always kind of chaotic and were definitely the most difficult and the most triggering of my symptoms. |
|  | Y2540: Relationships with others are, I think, one of my biggest motivators. Because I think all we ever really wanted was to have people around and you know, lots of relationships... I think it's just one of my biggest motivators, and also one of my biggest stressors, because when I put myself out there, I can't. I mean, if something, if anything goes off then I get triggered. |
| Theme 3: Crossroads and uneven change over time. |  |
| Subtheme 3a: Two roads diverged: recovery as the only option. | Y2097: I was also just really tired of my own crap. And it wasn't in the derogatory sense of like, oh, I'm so awful! I'm so stupid! More like, this sucks. I can't keep living my life like this, and then using that to fuel my motivation to get out. So rather than having all the stress weigh me down, it's like I'm using the stress to kind of push me back up out of whatever hole I dug myself into. |
|  | Y2575: I never thought I would get better, right? You think, you're diagnosed with BPD, and you're like, "Oh, I'm going to kill myself" like, that's how my life is going to end. Because that's what everybody does, right? And once I realized, okay. That's not what I'm going to do. That's not what I want to do. That's not what I envision doing. And I've got to find an alternative, right? Like I don't have another option. I have to find a way to live with this, and to survive this. |
|  | Y1930: I hit a point where I realized, if I keep doing this, I am going to die. Period... Or I can start doing things differently, and I can try to get better. There's no guarantee that that's gonna happen. But this, this path is definitely guaranteed. There's only one way this can go. And at this point I need to make a choice, either choose to just say, fuck it and let it happen, or I choose to be proactive and actually start doing something about it. |
| Subtheme 3b: Beyond behavior: the persistence of internal distress. | Y2143: I still deal with a lot of the symptoms internally, like I don't externalize them too much, but I'll still split over people – I think that's the term – pretty frequently... I still get stuck in that a lot. But I don't really get angry externally. For example, I don't really have the explosive episodes, yelling, whatever, at people that I used to, 'cause I can control it pretty well now. |
|  | Y2327: The areas I've struggled with more and have – I won't say have gotten worse, I don't think anything's gotten worse, but have definitely been slower to recover, and have been more of an up and down journey – have been again, my own self viewpoint and my relationship with others as a result. And then I've just, I've struggled with self-harm for a long time, and I don't necessarily do it anymore. But I find myself kind of going back to that mindset where I want to, and it's been a very up and down journey with that, so it's been a little difficult. But I mean it hasn't gotten worse. And again, I haven't done it in a very long time, which is still an improvement. It's just very hard to kick that mentality that gets you to the point of doing it. |
|  | Y2393: I feel like there's more of a heavier focus on...let's try to fit you in the societal standard mold as soon as possible, so you can be a functioning adult and human. But then you get there and you're left with the remnants of ... all of the thoughts that led me to do those actions to begin with. |

**Figure 1.**
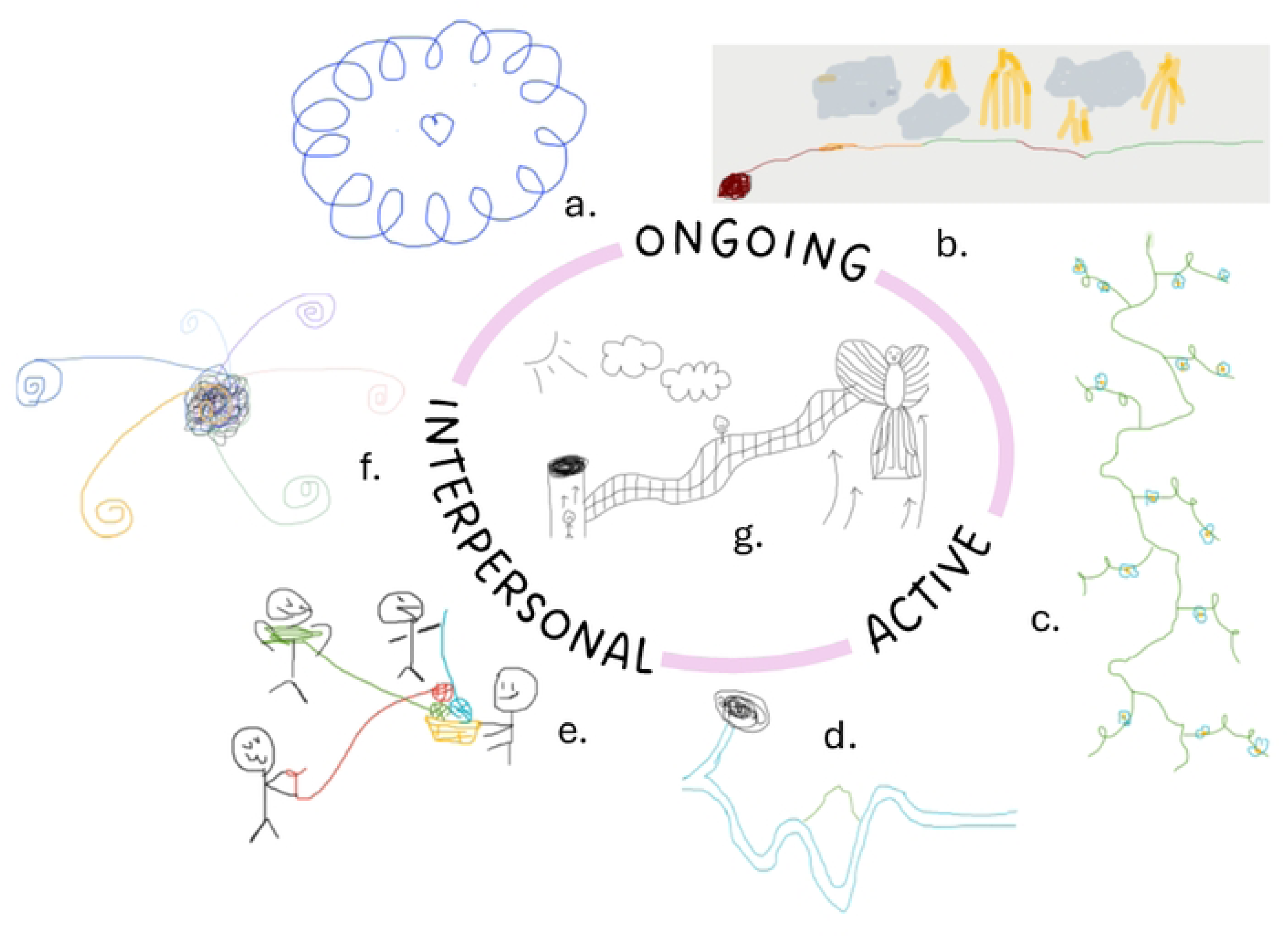
Drawings of the recovery experience. Participants made quick sketches of “what recovery looks like” during their interview visits, yielding drawings illustrating theme 2, the nature of recovery, and comments to explain the drawings. Representative drawings are shown illustrating subtheme 2.1 The ongoing nature of recovery (a: “…my journey is like, I don’t think there’s a beginning or an end. It’s like a cycle, like a circle, but there are like ups and downs in the circle, and there are times where I feel like I’m moving forward and times where I feel like I’m moving backwards, but it’s not like moving backwards is necessarily bad, because it’s a circle…. And now I have better tools to not suffer in this circle”, b: “It started very much like a hole, and then red, yellow, green, because it was rough, then it was okay, then it was good. And then it’s not linear… there were times where it was cloudy, hazy, like when you‘re relearning… almost like you’re a kid, you’re relearning what’s the appropriate way to react when something happens. So it’s cloudly, like almost unclear, like who am I if I am completely changing? … And then I tried to put a little bit of sunbeams through, because I feel like sometimes, especially compared to before, I’m like wow, this is a very serene way of living. But again sometimes it’s cloudy”), and subtheme 2.2 the active nature of recovery (c: “My journey, it’s very up and down, crazy… times where I’m doing good, times where I have really steep falls, and I’m in a really dark place. But then I’ll get better again, get a little bit more stable. …I like to draw it as like a flower vine to remind myself that in nature, nothing’s perfect, and there’s some beauty in things, and you never know what’s gonna happen,” d: “I realized that I had two paths. One was into this mysterious, unknown direction, and one was straight screeching into a black pit of death. So that’s my upwards path. And then the downwards path is winding. It’s hilly. It’s difficult. Sometimes it’s easier going downhill. Sometimes it’s really steep. But then, ultimately, at the end, it kind of levels out”) and subtheme 2.3 the interpersonal nature of recovery (e: “So we have me in the middle and I’m holding a little yarn basket with little balls of yarn. And each ball of yarn has a little thread that goes out to people …to represent my friends and my partner… Since realizing that I have BPD, [I’ve been] trying to make sure I’m prioritizing other relationships as well and not just my partner… So, you know, he still has all his green thread because we’ve been together for a long time. We’re very important to each other. But I’m also trying to make sure that I have other people with their own little yarn balls,” f: “I’m in the center and that’s my thought process, my emotions, my everything. They’re…all over the place, but they’re also very influenced by outside people. Each different swirl is a different person who is close to me or not close to me, and then the yellow one is a thicker line because they have a heavier influence on my thought process and on my emotions. …my thought process in the center is made-up of black, myself, but also made-up of all the different influences from the other people.”) Many drawings included elements of several aspects of recovery, as in g (“Being in a place where I saw no hope at all, I was convinced that I would end up taking my own life. I had no idea where to start in terms of addressing the struggles that I was living with and that I imagined for myself. And coming out of that desperation, through addressing my symptoms, working with my providers, having the support of my family and my closest friends, working towards becoming a peer support specialist, and feeling like I had and have a purpose in this life to help others establish and maintain recovery in that role…That’s supposed to be a phoenix. Even though it looks like a butterfly. And then I’m now, I think, in the middle of my journey towards becoming a phoenix. And I might always be in the middle of that journey. But I’m not in the hole. And I hope I don’t ever go back there.”

## 1. Differing attitudes toward language

To understand the impact of language, we asked participants to discuss their views on the term “recovery” and how the BPD diagnostic label shaped this understanding. We identified three subthemes: a) labels offer a useful framework, b) labels risk contributing to stigma, and c) an underlying shared sense of recovery despite varied language.

Vocabulary preferences varied within the sample; for clarity, we use the term “recovery” as a broad label to refer to the process of positive change over time.

### 1A. Shared vocabulary: labels offer a framework for self and other

Participants found labels such as “borderline personality disorder” and “recovery” useful both internally and externally to describe these experiences to others. Many participants described that having a diagnostic label validated their internal experiences. The label BPD framed “frustrating” patterns and experiences as “a symptom of sickness”, rather than personal failures that participants “couldn’t fix” (Y2088, Y2097). Participants reported feeling “happy to get the diagnosis” because it gave them a label as to why they had been struggling, “giving a name to [their] experiences” (Y1697, Y1612).

The term “recovery” was similarly useful to participants for making sense of their own experiences, giving participants language for feeling “a lot better,” and “managing well” (Y1832). For Y2088, “recovery” helped them understand and acknowledge “yes, I do have these issues and I need help.” Some participants felt particularly strongly about the specific verbiage: Y2575 preferred the word recovery to be capitalized to emphasize that it “should be taken very seriously.”

Participants also found the diagnostic label useful for connecting with other people. Equipped with a diagnostic label, participants could reach out to people close to them to open discussions and gain support (Y2401, Y2292). The label also enabled participants to seek support and community in online forums and support groups, connecting with people experiencing similar difficulties. Relating to others in these settings, connected by a shared vocabulary, allowed Y1612 to feel less “wrong and alone” and helped them cope. Participants generally felt that recovery in BPD merited greater public awareness: Y2097 noted that in the classroom and in casual conversations, “no one talks about what recovery…looks like” for people with BPD, with psychoeducation focusing instead on symptoms and experiences of “dealing with” people with BPD. They wished that there would be more use of the word “recovery,” inviting further discussion about the possibility of recovery from BPD.

### 1B. “I’m not broken”: stigma and black and white thinking

In contrast to the previous section, some participants had a negative response to the term “recovery,” finding the term stigmatizing and inaccurate. Having to “recover” from BPD implied to these participants that they were “broken” (Y2634). Even within the current study context, Y1330 “cring[ed]” at the word “recovery” prior to their interview with the research team, remarking “that’s gross,” because “I don’t think I’m broken,” as they felt the word implied). Y1612 preferred to use the phrase “in treatment for” because “recovery” gave the implication that “there’s something wrong with you… I don’t think there’s anything wrong with me.” Y1832 felt similarly and preferred “friendlier” words such as “living a meaningful life” or “managing symptoms.” To some, “recovery” was inaccurate, implying that symptoms would go away completely and that they would be cured (Y2692, Y2143). Improvement from BPD was considered asymptotic; while the goal was continual improvement, participants across the board acknowledged that they would never be fully better. Y1330 mused:

> *even after I graduate DBT I [won’t] consider myself recovered from BPD. I think it’ll probably always be like a part of who I am. Like I might not have as many symptoms, or it might not affect me as deeply. I think it’ll always be part of who I am… I’m a person with BPD.*

Y1832 succinctly outlined their resistance to the term recovery: “this is who I am forever.”

### 1C. Beyond labels: negotiating meaning and language of recovery

We specifically asked for participant perceptions of the terms “recovery” and “remission,” two common terms used to describe a stable positive change in people with BPD [33]. Specific vocabulary preferences varied, as outlined above, and at times were directly contradictory, as further outlined below. For example, Y2094 preferred remission to recovery because they found the implied medical connection validating:

> *With remission, what comes to mind are physical health illnesses, almost like cancer, because that word is used a lot in that setting. So I like that… I like the medical side of it and viewing mental health illnesses more as a medical thing.*

Directly contradicting this preference, Y2253 disliked the term remission, finding the appropriation of medical terminology to be “diminishing things, like people who have cancer and actually have gone into remission.”

Several participants, meanwhile, found the language of “recovery” and “remission” to be interchangeable. Recovery and remission were at times described to be variants of the same concept, for example Y1938 outlined “it’s the same thing, of course. I think if you want to talk about recovery, it specifically identifies what you’re gonna *do*. I guess it’s a *verb for remission*” [emphasis added]. Y2045 found little value in distinguishing the terms: “I’m like neutral now […] It’s like whatever’s helpful to talk about the thing and whatever other people use, like I’ll use it, so yeah.” Many participants shared that they had not considered the term “recovery” and had their own vocabulary that they preferred, even after being provided with this term. Others simply used recovery because it was more widely known, and therefore more easily understood by those around them (Y1832, Y3415).

## 2. Recovery as a dynamic and relational process

The second theme that we identified related to the nature of recovery. Regardless of whether they ultimately preferred “recovery,” “remission,” or some alternate language such as “healing” or “managing symptoms”, participants rejected language that implied an expectation of being “cured,” preferring language that emphasized a common understanding of the nature of recovery as a process that is a) ongoing, b) active, and c) centrally interpersonal.

### 2A. “A lifelong journey”: recovery as a process, not an endpoint

Participants viewed recovery as an ongoing process rather than a binary state of being. For instance, Y2634 viewed recovery as “doing better than you were before,” not a “yes or no” or “black or white.” Words denoting permanence and longevity such as “lifelong,” “long,” and “forever” were frequently brought up, referring to recovery as a “lifelong journey” and “lifelong battle.” Y1832 expressed hesitation about the possibility of being free of BPD, stating that they will “forever be managing this [disorder] and just being in recovery.”

Notably, participant drawings illustrated the course of recovery using a variety of symbols and yet seemed to converge on its nonlinear nature. As illustrated in Figure 1 drawing A, Y1330 described their recovery journey as a “cycle” characterized by “ups and downs,” where at times they reported feeling like they were both “moving forward” and “moving backwards.” Similar language figured prominently in interviews to characterize the recovery process, such as “ebbing and flowing,” “dynamic,” and “fluctuating.” Several participants accepted setbacks and difficult periods as part of the process of recovery. Y1932 embraced the possibility of lapses in their recovery: “I try my best… I cannot guarantee that I’m not gonna blow up one day for something stupid.” Y2138 echoed similar sentiments: “I feel like even if you’re in recovery, you feel like you’re recovered, etcetera, you can still go back to the behaviors. You can still have a slip up.”

### 2B. My recovery, my responsibility: embracing agency

Many participants emphasized recovery as an active process—not only due to its dynamic nature, but also in the sense that they were active agents in their recovery journey. Y2088 described how discussing the diagnosis with their therapist led to “an understanding that it’s not my fault that I’m like this, but it’s my responsibility to deal with that.” Y1930 noted the active engagement and intentionality involved in recovery, describing it as “more of a verb than a thing that happens to you… it’s a doing and it’s a process.” Y2080 contrasted their view of recovery with the more passive role they associate with being in treatment:

> *Being in recovery is a more active and involved process than just being in treatment… treatment to me, means, like… you’re going to therapy, you’re showing up… being in recovery is like, you’re actively doing the work to get better*.

Participants generally agreed that recovery requires continuous effort and the constant use of skills they’ve acquired to manage symptoms. Y2649 equated recovery with “ongoing work,” which they describe more closely aligns with their experience of BPD, whereas the term “cured” would seem “miraculous” and diminish the “hard work” they have put in. Specifically, participants characterized their recovery as sustained, everyday work, referring to it as managed on a “day-to-day basis” (Y2350) and requiring “daily maintenance” (Y1930).

### 2C. Healing and harm: connection as both struggle and goal

The strongest point of convergence across participant interviews was the primacy of interpersonal experiences in BPD recovery. For instance, improved relationships, particularly within the context of family, friends, and romantic partners, were one of the most widely cited goals for recovery. For Y2692, recovery would entail feeling more secure in their romantic relationships:

> *I wouldn’t feel like my world is falling apart if my partner went out for the night, like, you know, being able to deal with absences, short term absences in your life, like a normal person. That’s what recovery would be for me*.

Several participants also expressed a desire to maintain a stronger sense of self within their relationships. For instance, Y1612 expressed the goal of relying less on others and reclaiming their emotional well-being. Y2655 highlighted the unique challenge romantic relationships pose to their recovery, asking “Why am I able to be so successful in all these other areas of my life but I’m not able to be successful in romantic relationships?”

In centering on relationships as both a desired area of change and core source of distress, participant narratives highlight the complexity of recovery—while improved relationships were viewed as a valued aspect of recovery, the pursuit of that closeness was often feared and experienced as destabilizing. Y2540 captured this by characterizing their relationships with others as one of their “biggest motivators” yet also one of their “biggest stressors.” Y2506 described a push-pull dynamic of desiring yet avoiding connection to others:

> *[Interpersonal relationships] give me a lot of hope and motivation and support, and they’re a huge goal. But they’re also really hard and scary and difficult and tiring to manage… it’s very scary to be vulnerable…it feels sometimes easier to avoid.*

Establishing supportive relationships, both platonic and romantic, was reported as an important aspect of recovery. The capacity for mutually supportive intimate relationships was emphasized by Y1938, who stated that “being in a relationship with someone I fully love and trust, that I feel loves and trusts me, really, really, helps. That’s a big factor.” Other participants focused more on a broader social system, including others with similar experiences of BPD and mental health challenges. These relationships, offering a powerful sense of validation and understanding, were described as particularly meaningful. Y1330 reflected on the affirming experience of having friends who share similar struggles:

> *For the majority of my life, I have not felt understood… Knowing that my experiences actually aren’t unique, and I’m not the first person and I won’t be the last person to experience them, and not feeling alone, and not feeling different… feeling [a] profound sense of belonging with the people I surround myself with, I think, has helped immensely.*

## 3. Crossroads and uneven change over time

The final domain category analyzed the chronological development of recovery, delving deeper into what factors impact the ups and downs of the recovery journey. Two salient subthemes were identified when characterizing the “typical” recovery timeline: a) a decision point leading to seeking a higher level of care and b) common patterns of symptom remission.

### 3A. Two roads diverged: recovery as the only option

Receiving a diagnosis was often described as a critical point for realizing that recovery was one of only two options. Y1330 described the decision point that led to their determination to find treatment:

> *I was just so miserable, and I knew there had to be another way, and I was like ‘No.’ I was like, I can’t live like this anymore. I literally knew I couldn’t, I knew I was gonna kill myself if I continued living that way. And so I was like, I need to give everything to see if this works.*

Several other participants echoed this sentiment, highlighting seemingly rock-bottom points in their own lives wherein they realized that they had no choice but to make an active attempt at recovery. This pattern also appeared in some visual representations of recovery. When asked to draw their personal understanding of recovery **(Fig 1D)**, Y1930 described their recovery drawing:

> *I realized that I had two paths. One was into this mysterious, unknown direction, and one was straight screeching into a black pit of death. So that’s my upwards path. And then the downwards path is winding. It’s hilly. It’s difficult. Sometimes it’s easier going downhill. Sometimes it’s really steep. But then, ultimately, at the end, it kind of levels out.*

Hospitalizations played an ambivalent role in early recovery experiences: “a lot of the hospitalizations were very unhelpful. And actually, were more traumatizing. Which is unfortunate. *They kept me alive, but they left me with more trauma*” (Y2350, emphasis added). Many participants acknowledged that despite the difficulty, hospitalizations ultimately played a critical role in their recovery. In the context of a paucity of treatment options, Y2059 reflected that “the only way in the past that I’ve been able to get additional support when I needed it was being hospitalized.” This sentiment was echoed by several participants (Y2143, Y2253, Y2350, Y2327, Y1594). Hospitalization appeared throughout the interviews as a sort of “necessary evil,” an experience always described as negative. However, participants often reflected back on hospitalizations as critical juncture when they understood the severity of their struggles and set on a path to recovery. Importantly, reported positive outcomes of hospitalization often fit into the larger pattern of participants who reported a personal decision point within the process of receiving a diagnosis or seeking treatment: a higher level of care was helpful insofar as it guided participants to the decision to take recovery seriously. Ultimately, whether a result of hearing the diagnosis, being hospitalized, or some other internal pressure, this experience of taking ownership over one’s recovery experience was reiterated by participants as a critical point in the recovery journey.

### 3B. Beyond behavior: the persistence of internal distress

The more behavioral components of BPD – non-suicidal self-injury (NSSI), risky impulsive behaviors, intense interpersonal conflicts – were often reported to be easier to “eliminate” than difficult cognitive patterns – fear of abandonment, intense emotional highs and lows, suicidal ideation (Y2310). While the urgency of addressing more severe behavioral distress is obvious, participants suggested that the core internalizing symptoms remained pervasive across stages of the recovery journey, leading to ongoing distress even if functional impairment had been reduced with treatment. Y2393 reflected on the complexity of their experience of BPD, noting a gap after years of treatment between outward signs of progress and ongoing internal distress:

> *Recovery for me with my borderline is the decrease in the outward compulsions or symptoms… I have not really felt an actual internal, like, dramatic decrease in the amount of intrusive horrible thoughts that’s happening… I can’t get the bad thoughts to shut up.*

Many participants viewed this amelioration of internal symptoms as the final step of the recovery process, the distinction between “recovery” and “*full* recovery”. Participants described full recovery largely in relation to their internal cognitive experiences: an absence of “impulses” for “destructive behaviors” (Y1330), “just being happy and not having the kinds of thought patterns or obsessive behaviors that I have right now” (Y2045). A sense of stability and influence over their emotional experiences remained a goal even for those who have improved their social functioning. Y2292 described how even as they have maintained a stable job and happy relationship for over four years, they still would love to feel “more in control when there are things that make me really, emotionally dysregulated”

Y2292 suggested a more dimensional understanding of recovery, stating that “recovery shouldn’t be based on the number of symptoms but more on severity of impairment.” Importantly, participants were quick to identify that they didn’t hope for a *full* abatement of intense cognitive/emotional experiences. For many participants, their intense emotions were a crucial, and often positive, piece of their identity, and while they hoped for a lessening of dysfunction, they were similarly distressed at the concept of emotional blunting. When asked to imagine full recovery, Y3415 paused and reflected:

> *My first answer was going to be that you’re more or less indistinguishable from the average person, but I don’t know. I left my BPD diagnosis behind a couple of years ago, and I feel like I want to be as far away from average as possible. So I don’t know.*

## Discussion

This study explored how individuals with lived experience of BPD understand recovery, with particular attention to the language used to describe change, the defining features of recovery, and the factors shaping its course over time. This work was motivated in part by the persistent discrepancy in the literature noted above: long-term studies consistently document substantial rates of symptomatic remission, yet recovery, as lived and understood by individuals themselves, remains more difficult to define, measure, and predict [8, 32, 33, 41, 42]. Participant accounts help clarify this gap, suggesting that remission and recovery may reflect related but distinct dimensions of change.

Drawing on a relatively large qualitative sample (n = 41), including individuals across a wide range of symptom severities, treatment histories, and stages of recovery, this study extends prior qualitative work that has often relied on data from smaller and more homogeneous clinical samples [14, 22, 26, 34]. The inclusion of men and non-binary/genderqueer gender identities also broadens a literature that has frequently focused exclusively on women’s experiences.

### Language and meaning-making in recovery (Theme 1)

The first theme identified in this work concerned the multifaceted role of language in shaping how recovery is understood. Consistent with prior studies, some participants in this cohort described the term “recovery” as imperfect or uncomfortable, particularly when it was taken to imply cure or a return to a premorbid self [14, 38]. However, for others, the diagnostic label BPD or the term recovery provided a framework for understanding previously confusing experiences and communicating with others. Participants described feeling validated by “giving a name” to their experiences and being able to locate communities of others with similar struggles. However, the same language felt stigmatizing or overly deterministic to some of those interviewed. Extensive literature documents stigma associated with BPD diagnoses [43, 44], but our findings suggest that the effects of language are not limited to stigma. Language also shapes how individuals imagine their futures, including whether recovery feels possible or coherent.

Another finding that emerged across interviews was that disagreements about terminology often masked deeper agreement about the underlying process of change (Theme 1C, e.g., “it’s the same thing, of course…”). This suggests that debates about terminology may sometimes obscure a shared experiential core. Clinically, this points to the importance of exploring patients’ own language and definitions of progress rather than assuming that standard terminology will be meaningful or helpful [45].

### The nature of recovery: process, agency, and relationships

The second theme concerned how participants understood the nature of recovery itself, specifically recovery as a journey (rather than a binary state), the importance of agency in recovery, and interpersonal relationships as integral to recovery. These accounts are consistent with broader conceptualizations of personal recovery in mental health [46] and with prior qualitative work in BPD [16, 38].

Of interest, we identified a point of tension in discussion of the role of agency in recovery in that it was often paired with talk about not being at fault for having BPD. The philosopher Hannah Pickard has framed this tension in a conceptualization of “responsibility without blame” [30, 47], a framing that mirrors the emphasis on both change and validation in Dialectical Behavior Therapy.

Perhaps the most consistent finding across interviews was the centrality of interpersonal relationships. Participants described improved relationships, greater emotional independence, and a stronger sense of self within relationships as key markers of recovery. Relationships were also frequently described as sources of distress and vulnerability, creating a push–pull dynamic in which connection was both desired and feared. Some described improved stability by avoiding interpersonal stressors or limiting close relationships, at least temporarily. While this strategy sometimes reduced crises, it did not necessarily resolve underlying fears or vulnerabilities. These findings are consistent with the formulation of interpersonal interactions as a central concern in BPD [48]. They also fit with prior qualitative research that identifies social connections as both supportive of the recovery process and as a hoped-for goal [12, 14, 22, 34, 38].

### The shape of recovery and uneven change over time

The third theme focused on how recovery unfolds over time. Most important in these accounts was not simply receiving treatment but deciding to actively engage with recovery. Participants also described uneven progress across symptom domains, with internal distress often persisting beyond behavioral stabilization. These accounts parallel longitudinal findings indicating that some symptoms remit more quickly than others and that recovery may involve continued internal struggle even in the context of improved functioning [33, 49]. Participants often described relief from internal distress as the most difficult and most meaningful phase of recovery, suggesting that definitions based primarily on behavioral or functional criteria may overlook an important dimension of lived experience. Findings from our diverse sample support and expand upon previous qualitative research conducted in smaller, more homogeneous samples, integrating perspectives from individuals who were highly symptomatic and those who had been in recovery for years, some actively in treatment and others not. Previous literature primarily emphasized specific symptom change – i.e. what does a person “recovered” from BPD look like? Our diverse sample of participants expanded upon this picture, outlining the shape of recovery from the initial decision to change through to a more stable phase of daily maintenance.

### Integrating contradiction and dialectical understanding

Across themes, participants frequently expressed seemingly contradictory views: valuing and resenting diagnoses, seeking closeness while fearing it, and describing themselves as both improved and still struggling. Rather than appearing inconsistent, these accounts often reflected a more nuanced and integrated understanding of experience. This pattern aligns with clinical approaches such as dialectical behavior therapy and mentalization-based therapy, which emphasize the capacity to hold multiple perspectives and tolerate ambiguity [50, 51]. Recovery, in this sense, may involve not only reducing symptoms but also developing greater psychological flexibility and tolerance for complexity.

### Strengths and limitations

This study has several strengths. The sample size is large for qualitative interview research, and recruitment outside a single clinical setting allowed inclusion of participants with diverse durations of BPD, treatment histories, and levels of current symptom severity. People with more recent onset and treatment of BPD may be able to reflect with more detailed recall of those experiences, whereas those with a longer history of BPD and longer recovery journey may be better able to describe an overall trajectory of recovery. Inclusion of a wide range of participants strengthened the nuance of these aspects of the report. The sample also included greater gender diversity than many prior qualitative studies of BPD. Interviews were piloted with an individual with lived experience of BPD, strengthening the relevance and sensitivity of the interview guide.

Several limitations should be noted. For instance, the sample was limited to English-speaking adults in the United States. Cultural and systemic differences may influence how recovery is understood and discussed. Although the sample was diverse in several respects, it cannot fully represent the experiences of individuals from all racial, cultural, or socioeconomic backgrounds.

## Conclusion

Participants in this study described recovery in BPD not as a discrete outcome but as an evolving process shaped by language, relationships, and personal agency. Their accounts help clarify why recovery remains difficult to define using narrow or binary criteria and suggest that incorporating lived-experience perspectives may help bridge the gap between clinical definitions of remission and individuals’ own understandings of meaningful change.

## Data Availability

Consistent with standards for qualitative research, we provided a table with representative quotes and full data will be made available upon reasonable request from members of scientific community.

## Acknowledgements

We thank Jonah Goldman and Giana Pittaro for their valuable contributions to data collection, as well as Asa Nelson-Ulsh, for his thoughtful insight on participants’ visual narratives. We are grateful to our participants whose narratives form the heart of this work. This paper is dedicated to honoring them and representing them with care, humility, and respect. Their courage, vulnerability, and generosity in sharing their experiences made it possible to explore these deeply personal and complex topics.

## Credit statement

**Verenice Torres:** Conceptualization, Methodology, Formal Analysis, Investigation, Data Curation, Writing – Original Draft, Writing – Review & Editing, Project Administration.

**Thea M. McAfee:** Conceptualization, Methodology, Formal Analysis, Investigation, Data Curation, Writing – Original Draft, Writing – Review & Editing, Project Administration.

**Paulina Tein:** Formal Analysis, Data Curation, Writing – Original Draft, Writing – Review & Editing.

**Alexandra A. Alario:** Formal Analysis, Data Curation, Writing – Original Draft, Writing – Review & Editing.

**Wenjing Luo:** Data collection, Data Curation, Writing – Review & Editing

**Kaylin Foss:** Writing -- Review & Editing

**Sarah K. Fineberg:** Conceptualization, Methodology, Writing – Review & Editing, Supervision, Project Administration, Funding Acquisition.

**Sharon M. Nelson:** Methodology, Resources, Writing – Review & Editing, Supervision.

## Funding

This work was supported in part by the Brain and Behavior Research Foundation (Young Investigator grant 30865 to SKF) and the National Institute of Mental Health (NIMH) of the National Institutes of Health under award number 1K23MH123760 (to SKF). The work described in this article was funded in part by the State of Connecticut, Department of Mental Health and Addiction Services, but this publication does not express the views of the Department of Mental Health and Addiction Services, the State of Connecticut or the NIH. The views and opinions expressed are those of the authors.

## Disclosures

SKF provides ad hoc consulting services to Boehringer Ingelheim GmBH and serves on a Scientific Advisory Board for Oryzon. The other authors have no relationships to disclose.

## Notes

### Author Declarations

Yale Human Investigation Committee (HIC)

